# PRADclass: Multi-pronged Gleason grade-informed computational strategy identifies consensus biomarker features of prostate adenocarcinoma that predict aggressive cancer

**DOI:** 10.1101/2023.04.04.23288124

**Authors:** Alex Stanley Balraj, Sangeetha Muthamilselvan, Rachanaa Raja, Ashok Palaniappan

**Affiliations:** Department of Bioinformatics, School of Chemical and Biotechnology, SASTRA Deemed to be University, Thanjavur 613401. India

**Keywords:** Prostate adenocarcinoma, RNASeq transcriptomics, Gleason grading, Cancer aggressiveness, Differential gene expression, Monotonic expression, Pairwise contrasts, Linear modeling, WGCNA, Network reconstruction, Enrichment analysis, Grade-salient gene, Trait-specific key gene, Consensus root biomarker, Machine learning, Random forest, Risk stratification.

## Abstract

**Background:** Prostate adenocarcinoma (PRAD) is the most common cancer in men worldwide, yet gaps in our knowledge persist with respect to molecular bases of PRAD progression and aggression. It is largely an indolent cancer, asymptomatic at early stage, and slow-growing in most cases, but aggressive prostate cancers cause significant morbidity and mortality within five years. Automated methods to type the aggressiveness of PRAD are necessary and urgent for informed treatment management.

**Methods:** Based on TCGA transcriptomic data pertaining to PRAD and the associated clinical metadata, we used the grading guidelines of the International Society of Urological Pathology (ISUP), and converted the clinical information of a cancer sample to its Gleason grade. To model the distinction between aggressive prostate cancers (Gleason grade IV or V) and indolent prostate cancers (Gleason grade I or II), we performed: (i) Gleason-grade wise linear modeling, followed by five contrasts against controls and ten contrasts between grades; and (ii) Gleason-grade wise network modeling using weighted gene correlation network analysis (WGCNA). Consensus between the grade-salient genes from the statistical modeling and the trait-specific key genes from network modeling were used as features for learning a ternary classification: benign, indolent or aggressive malignancy.

**Results:** The statistical modeling yielded 77 Gleason grade-salient genes, viz. ten genes in grade-1, two genes in grade-II, one gene in grade-III, 34 genes in grade-IV, and 30 genes in grade-V. Using the WGCNA method, we reconstructed grade-specific networks, and defined trait-specific key genes in grade-wise significant modules. Consensus analysis yielded two genes in Grade 1 (SLC43A1, PHGR1), 26 genes in Grade 4 (LOC100128675, PPP1R3C, NECAB1, UBXN10, SERPINA5, CLU, RASL12, DGKG, FHL1, NCAM1), and seven genes in Grade 5 (CBX2, DPYS, FAM72B, SHCBP1, TMEM132A, TPX2, UBE2C). PRADclass, a RandomForest model trained on these 35 consensus biomarkers, yielded 100% cross-validation accuracy on the ternary classification problem.

**Conclusions:** Consensus of orthogonal computational strategies has yielded Gleason grade-specific biomarkers that are useful in pre-screening (cancer vs normal) as well as typing the aggressiveness of cancer. PRADclass has been deployed at: https://apalania.shinyapps.io/pradclass/ for scientific and non-commercial use.

## INTRODUCTION

Over the past decade, global cancer incidence has increased by 33%, affecting 16% of the aged and 13% of the growing population [1]. In 2020, 19.3 million new cancer cases and 10.0 million cancer deaths were reported [1]. Prostate adenocarcinoma (PRAD), more commonly prostate cancer, is the most common cancer diagnosis among men, and as serious a threat to men’s health as breast cancer to women’s health [2]. The rise in prostate cancer cases may be attributed to lifestyle changes, smoking history, diet, and environmental factors [3]. Prostate tumors are very heterogeneous histologically and clinically. Early-stage prostate cancer is largely asymptomatic and progresses slowly, and often has an indolent course that might take more than 20 years to impact the quality of life [4]. Some prostate cancers grow aggressively and cause significant morbidity and mortality within five years. Malignant prostate cancers might necessitate prostatectomy, with a lifetime of treatment-related disabilities [4, 5]. The aggressiveness of prostate cancer is a crucial factor when deciding treatment regimen [6], and detection of prostate cancer in early stages and continued active surveillance could be an advantageous management option [7].

Measurement of prostate-specific antigen (PSA) levels followed by Digital Rectal Examination (DRE) are conventionally used to diagnose prostate cancer, however this method is prone to overdiagnosis and does not stratify cancer as slow-growing (indolent) or aggressive (high-risk). Recognizing this key gap in medical diagnosis coupled with the intrinsic difficulties in manual histopathology, automated deep learning systems have been steadily evolving to stratify prostate cancer according to the Gleason grade [8]. Such methods work with tissue microarray or whole slide images of biopsies, and have been advanced as potential clinical aid to support reproducible pathologist grading [9–16]. Omics approaches could be useful in studying and characterizing the progression of prostate cancer and its conversion to an aggressive form [17,18]. Expression profiling (or transcriptomics) is useful to gain an understanding of the genetic factors of prostate cancer pathology [19]. Epigenetic processes also contribute to the development of prostate cancer. DNA hypermethylation of CpG-rich gene promoter regions is widespread in prostate neoplasia [20]. Transcriptomics and mutational status-based genomics of prostate tumors might enable the development of personalized therapeutics [21]. Following these cues, we have designed a study protocol for inferring the biomolecular foundation of the Gleason risk stratification of PRAD. Understanding and identifying Gleason grade-specific gene expression might be useful to achieving precise personalized management of prostate cancers. In this work, we have developed a multi-layered approach to address the above concerns to potentiate the differential diagnosis of aggressive prostate cancers (Gleason grades IV and V) versus indolent prostate cancers (Gleason grades I, II, and III).

## METHODS

The investigative workflow, presented in Figure 1, consisted of two major branches, statistical –omics analysis and network-based WGCNA analysis, which were integrated in the end to yield root biomarkers of Gleason grade-wise PRAD progression.

**Figure 1.**
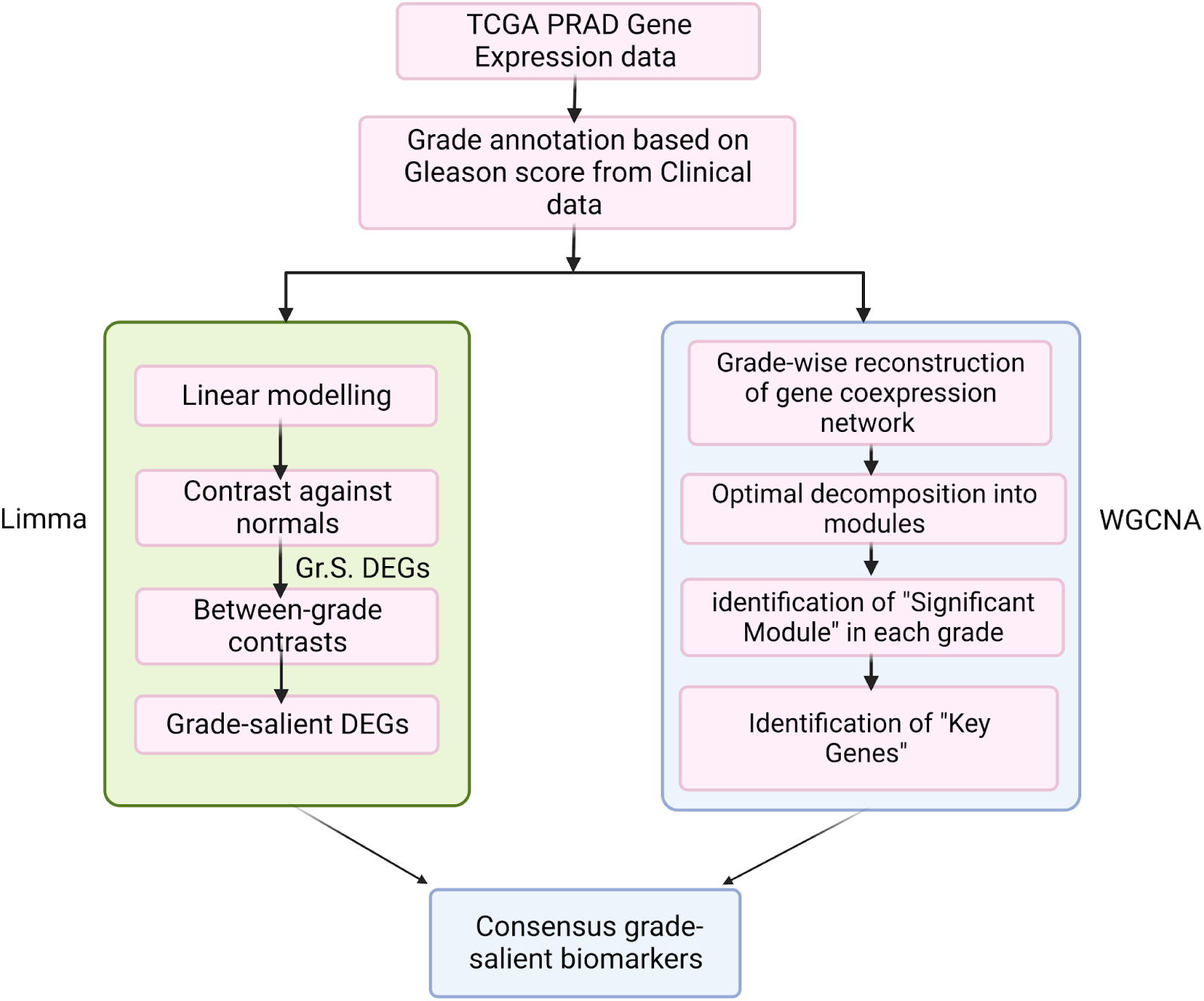
Workflow for identifying root biomarkers based on a consensus between statistical modeling and WGCNA analysis of PRAD transcriptome.

### Data acquisition and pre-processing

Public-domain TCGA datasets were used in our analysis [22]. RSEM-normalised PRAD RNASeq gene expression data was obtained from the firebrowse portal (gdac.broadinstitute.org_PRAD.Merge_rnaseqv2 illuminahiseq_rnaseqv2 unc_edu Level_3 RSEM_genes_normalized data.Level_3.2016012800.0.0.tar.gz) [23]. Associated clinical data was retrieved from the same portal (PRAD.Merge_Clinical. Level_1.2016012800.0.0tar.gz). Three Gleason grade-related clinical attributes were of interest:

1. primary Gleason pattern encoded as patient.stage_event.gleason_grading.primary_pattern
2. secondary Gleason pattern encoded as patient.stage_event.gleason_grading.secondary_pattern
3. Gleason score (sum of the primary and secondary patterns) encoded as patient.stage_event.gleason_grading.gleason_score

Information about the primary and secondary patterns is necessary to disambiguate a Gleason score of, say, 7, which could arise from either 3 + 4 or 4 + 3, with the latter having a worse prognosis according to ISUP guidelines [24]. Using these clinical attributes, the Gleason grade was derived (Table 1). The RNA-Seq data was matched with the clinical data using the sample ‘patient_bcr’ information. The genes with nominal variation in expression across samples (σ < 1) were removed. Samples with missing grade annotation (or ‘NA’) were also removed. All data pre-processing was done with R [25].

**Table 1:** Grading of prostate cancer samples, based on the Gleason pattern and ISUP guidelines.

| Gleason Pattern |  | Gleason Score | Derived Gleason Grade |
| --- | --- | --- | --- |
| Primary | Secondary |  |  |
| 3 | 3 | 6 | 1 |
| 3 | 4 | 7 | 2 |
| 4 | 3 | 7 | 3 |
| 3 or 4 or 5 | 5 or 4 or 3, respectively | 8 | 4 |
| 4 or 5 | 5 or $\geq 4$, respectively | 9 or 10 | 5 |

### Linear modeling with PRAD Grade

A grade-wise linear model of gene expression was constructed using the R *limma* package [26]:

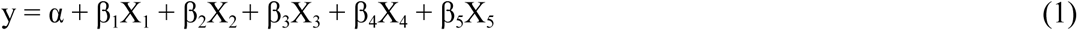

where the intercept α represents the baseline expression from control samples, and the β_i_ signify grade-wise log-fold change (lfc) in expression relative to control samples. Empirical Bayes adjustment [27] and correction for multiple hypothesis testing [28] were carried out to yield a grade-wise account of differentially expressed genes ranked by significance. This model facilitated the contrast between each grade and controls (Table 2), and genes with |lfc| < 2 in any grade were eliminated.

**Table 2:** Contrast matrix with control. Mean expression of samples in each grade are contrasted with the mean expression in control samples, to obtain grade-specific lfc with accompanying significance.

| Annotation | Grade 1 - Control | Grade 2 - Control | Grade 3 - Control | Grade 4 - Control | Grade 5 - Control |
| --- | --- | --- | --- | --- | --- |
| Control | -1 | -1 | -1 | -1 | -1 |
| Grade 1 | 1 | 0 | 0 | 0 | 0 |
| Grade 2 | 0 | 1 | 0 | 0 | 0 |
| Grade 3 | 0 | 0 | 1 | 0 | 0 |
| Grade 4 | 0 | 0 | 0 | 1 | 0 |
| Grade 5 | 0 | 0 | 0 | 0 | 1 |

### Pairwise contrasts modeling

To identify grade-salient genes, we modified the model of gene expression given by eqn. (1):

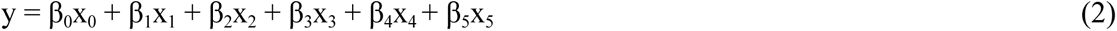

where the x_i_ are indicator variables encoding the sample grade, and β_i_ correspond to estimated weights for each grade [29]. The design matrix corresponding to these contrasts is shown in Table 3.

**Table 3:** Contrast matrix for executing between-grade contrasts using the `contrastsFit` function in *limma*. Five grades of cancer progression implied ^5^C_2_ possible pairwise contrasts, four for each grade.

| Derived annotation | Between-Grades contrast |  |  |  |  |  |  |  |  |  |
| --- | --- | --- | --- | --- | --- | --- | --- | --- | --- | --- |
|  | Grade1 - Grade 2 | Grade1 - Grade 3 | Grade1 - Grade 4 | Grade1 - Grade 5 | Grade 2 - Grade 3 | Grade 2 - Grade 4 | Grade 2 - Grade 5 | Grade 3 - Grade 4 | Grade 3 - Grade 5 | Grade 4 - Grade 5 |
| Grade 1 | 1 | 1 | 1 | 1 | 0 | 0 | 0 | 0 | 0 | 0 |
| Grade 2 | -1 | 0 | 0 | 0 | 1 | 1 | 1 | 0 | 0 | 0 |
| Grade 3 | 0 | -1 | 0 | 0 | -1 | 0 | 0 | 1 | 1 | 0 |
| Grade 4 | 0 | 0 | -1 | 0 | 0 | -1 | 0 | -1 | 0 | 1 |
| Grade 5 | 0 | 0 | 0 | -1 | 0 | 0 | -1 | 0 | -1 | -1 |

### Detection of grade-salient genes

Grade-salient genes were identified using a sequence of five filters:

i. A stringent adj. p-value (< 0.001) of the contrast against controls was applied to filter the significant differentially expressed genes. Such genes were assigned to the grade showing maximum |lfc|, producing grade-specific genes.
ii. (v) Significance of contrast with reference to other grades (adj. *P-*value < 0.05) was applied to the grade-specific genes, and those that pass all the four relevant contrasts (out of the ten possible pairwise contrasts) were identified as grade-salient genes.

### Monotonic expression

To ascertain any significance in the ordination of mean expression across grades, we used a model with PRAD grade (X) as a numeric variable:

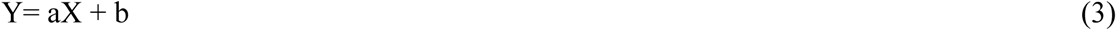

To identify genes whose expression is in step (monotonic) with PRAD progression, we checked for the following two patterns with respect to mean gene expression:

i. control < I < II < III < IV<V; signifying monotonic upregulation
ii. control > I > II > III > IV>V; signifying monotonic downregulation

### Gleason biomarker identification using WGCNA

The TCGA PRAD RNAseq data was normalized using voom and checked for outliers [30]. Genes were ranked based on the median absolute deviation in expression across the samples, and the top 5,000 genes were identified. The expression subset corresponding to these genes was annotated with the sample Gleason grade, and used to construct Gleason-grade specific co-expression networks using WGCNA [31], yielding five networks. For each such network, the smallest scale-free exponent with goodness-of-fit R^2^ > 0.9 was identified as the β value using WGCNA’s ‘pickSoftThreshold’. With β value fixed, we used the Pearson’s ρ to calculate the similarity matrix corresponding to each network, which was then converted sequentially into the adjacency matrix, topological overlap matrix (TOM) and dissimilarity matrix (given by 1-TOM). Hierarchical clustering was applied on the resulting dissimilarity matrix, and a dynamic tree cut algorithm with a cut height of 0.25 was used to partition the gene space into 20 co-expression modules.

The association between each module and the clinical trait of interest (namely, the Gleason grade) was finally analyzed to identify the modules functionally significant for PRAD progression. Based on the module-trait relationships, the following measures were computed:

i. gene significance (GS), defined as the Pearson’s ρ between gene expression and the trait of interest, i.e, Gleason grade (in other words, cor(gene_i_, Trait));
ii. module membership (MM), defined as the degree of association of a gene in a module with all the other genes in that module, i.e., the degree of internal connectivity of some gene in a given module;
iii. module significance (MS), defined as the mean of the unsigned GS of all genes in the module; and
iv. module eigengene (ME), defined as the first principal component of the module-specific gene expression matrix.

Based on the above definitions, among all the modules for a given grade, the one with the largest MS value was designated as the Significant Module of that grade. Among all the genes of a given Significant Module, those with module membership MM > 0.7 and abs(GS) > 0.5 were identified as trait-specific key genes. The trait-specific key genes were ranked by GS.

### Integrative analysis

The grade-salient genes from –omics analysis and trait-specific key genes from WGCNA were investigated, and their overlap designated as the Consensus Genes. KEGG [32] and Gene Ontology were used as reference databases for analyzing functional enrichment [33]. The outlier genes were visualized using volcano plots (-log_10_ transformed p-value vs. log fold-change), and the grade-salient DEGs were visualized using heatmap dendrograms. The novelty of the identified grade-salient genes was screened against the Cancer Gene Census v84 [34], Network of Cancer Genes [35], and the Clinical Trial Registry (www.clinicaltrials.gov). The top ten trait-specific key genes were used to reconstruct networks using STRINGdb [36].

### Machine learning model for high-risk aggressive prostate cancer

Gleason grade-specific biomarkers might make for good features in pre-screening benign hyperplasia from cancerous samples, and further discriminating between indolent and aggressive prostate cancers. Towards this end, a ternary outcome vector was encoded, differentiating among normal or benign samples, indolent PRAD samples (grades 1, 2, and 3) and aggressive PRAD samples (grades 4 and 5). Multiple machine learning algorithms were used to address the ternary classification problem at hand. Specifically, the consensus genes from integrative analysis were used as the feature space for model-building with Random Forest [37,38], SVM [39], neural networks and XGBoost classifiers [40]. The pre-processed dataset described in the preceding Methods was used for model training, and k-fold cross-validation (k=10) was done to optimize model hyperparameters. Post hyperparameter-optimization, the classifiers were evaluated using average performance on all the folds, and the best-performing model identified. This model was re-built using the full dataset and deployed as an R Shiny app [41].

## RESULTS

The gene expression matrix consisted of 20,531 genes x 551 samples, with 52 controls. After preprocessing we obtained a final dataset of 18,327 genes x 538 samples, with the following distribution: 50, 149, 95, 70 and 123 samples in ISUP-based I, II, III, IV and V Gleason-grades, respectively. The dataset is available as Supplementary File S1.

The dataset was log_2_-transformed and observation-weighted through voom using the *limma* package. The linear model (eqn. 1) yielded 15257 significant genes (adj. *P* < 0.05). A more stringent significance cutoff (adj. *P* < 1E-05) yielded 7965 significant genes. The top ten genes of linear modeling, ranked by adj. *P*-value, are all upregulated, and their lfc with respect to control samples are shown in Table 4. The detailed results of linear modeling for all genes are provided in Supplementary File S2. Fig. 2 presents the expression patterns of the top ten differentially expressed genes from linear modeling. Principal components extracted from the expression patterns of the top 100 genes were effective in discriminating cancer samples from control samples (Figure 3a), in contrast to the principal components of the expression patterns of a random subset of 100 genes that failed to demonstrate any such separation (Figure 3b). Violin plots of all the top 200 genes could be found in Supplementary File S3.

**Figure 2.**
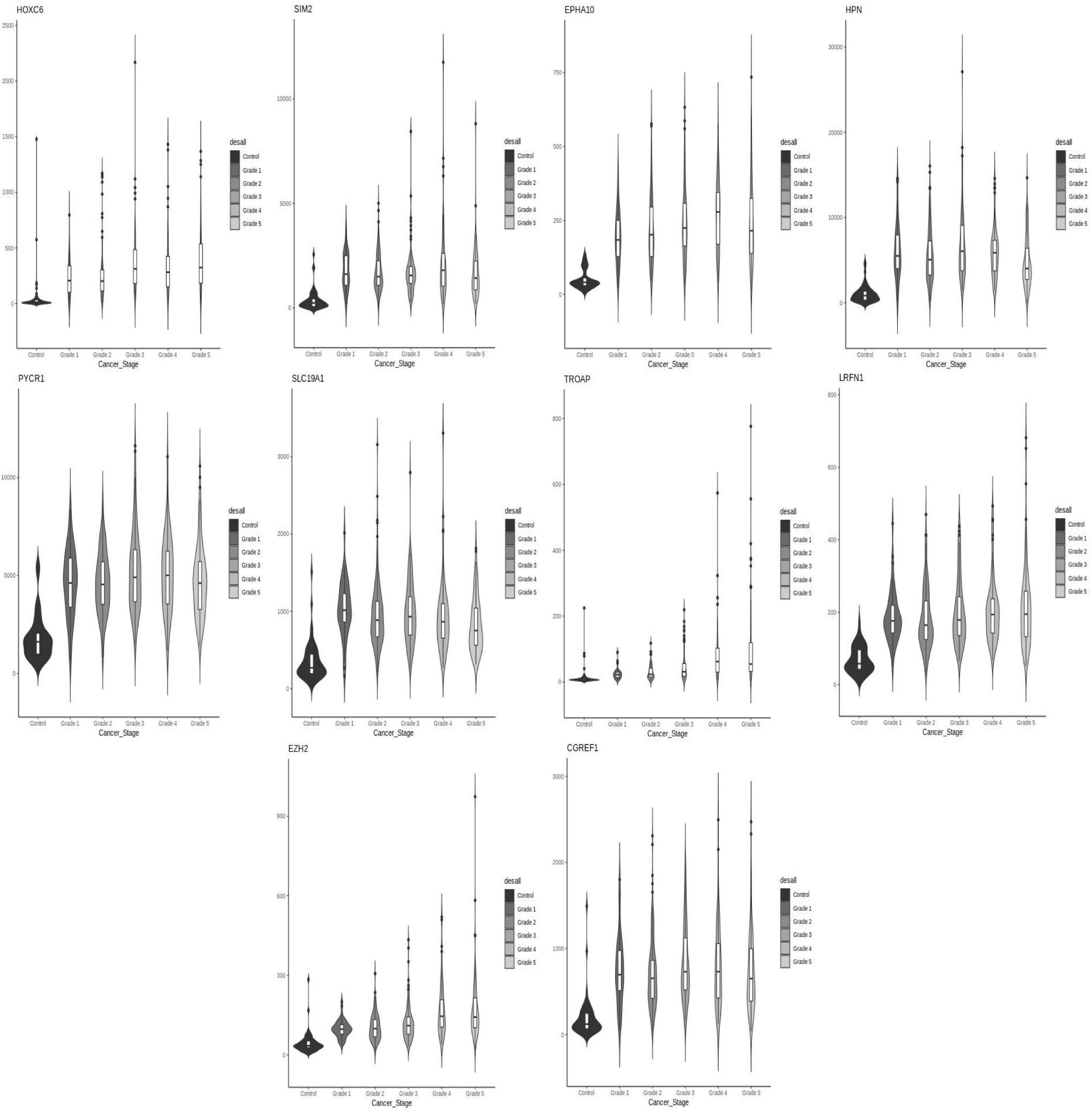
Violin plots of top 10 linear model genes. For each gene, notice that the trend in expression could be either upregulation or downregulation relative to the control. All the top ten genes are upregulated. It could be seen that the linear trend does not enforce lfc to attain a maximum in grade-5.

**Figure 3.**
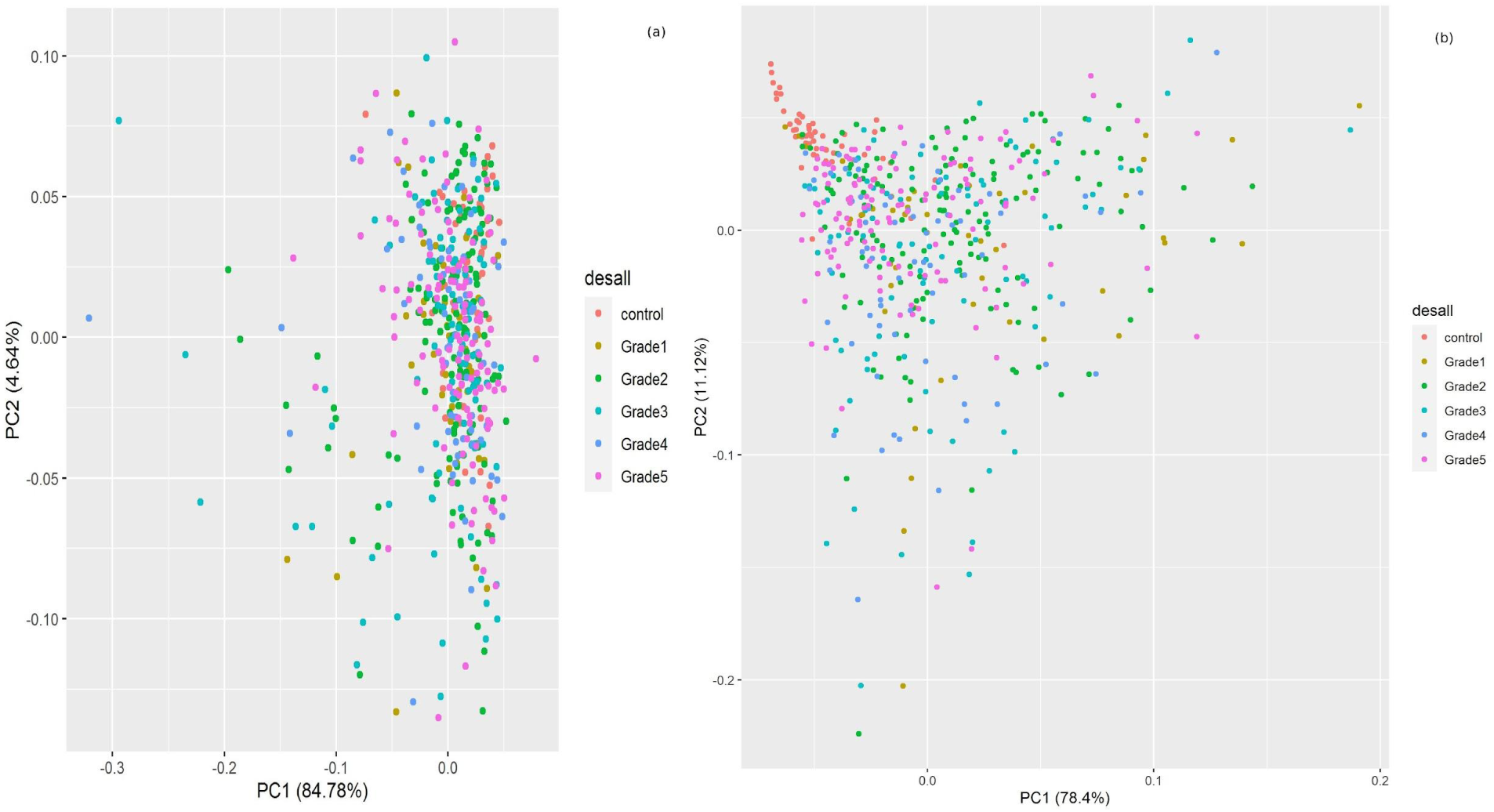
Principal components analysis of cancer vs control. (A) The first two principal components of the top 100 genes from linear modeling are plotted. It could be seen that control samples (red) clustered independent of the cancer samples (colored by grade). (B) The same analysis repeated with 100 random genes failed to effect a clustering of the control samples relative to the cancer samples.

**Table 4:**
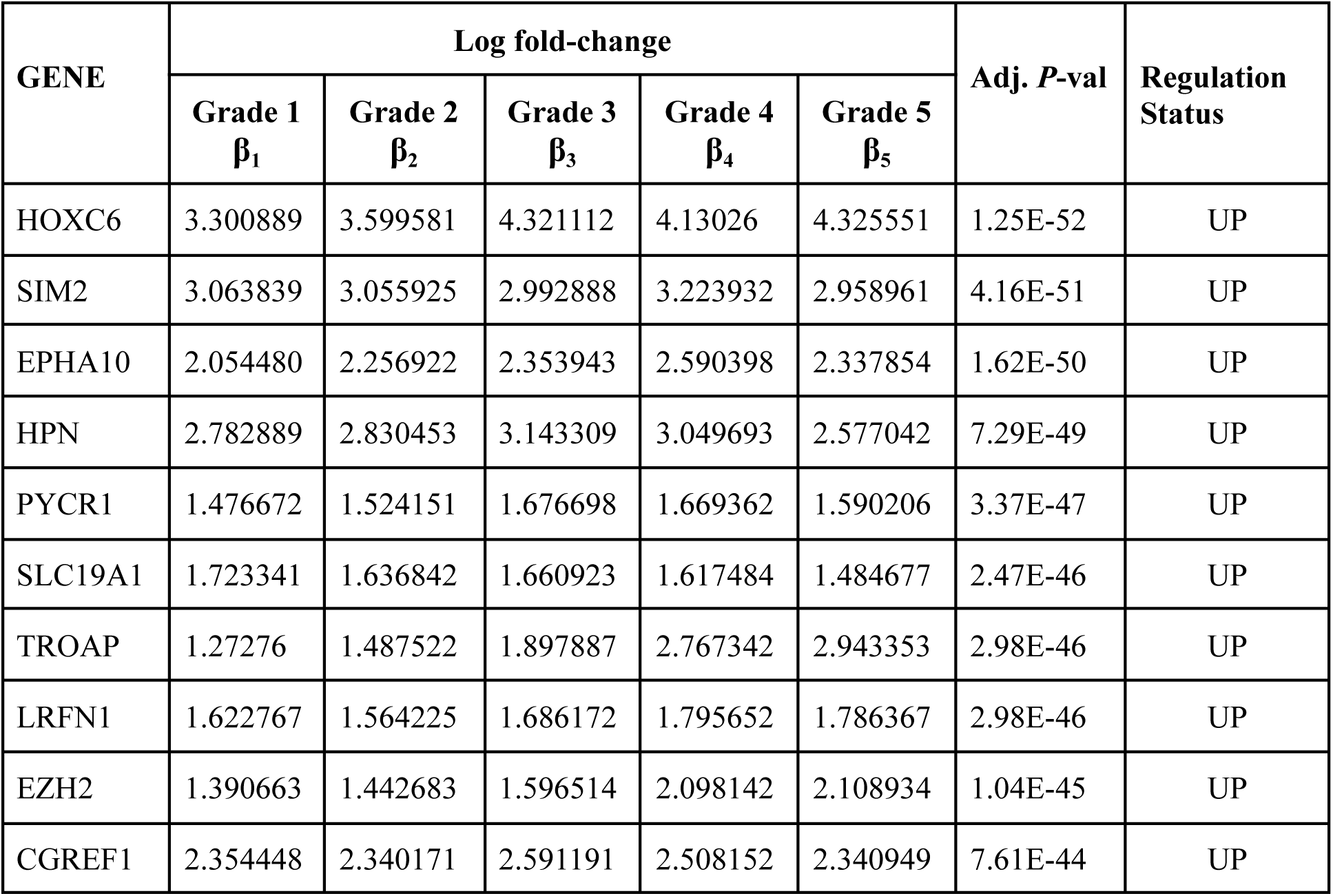
Top ten genes of the linear model, ranked by significance. The lfc of mean gene expression in each grade relative to control samples is given, followed by adj. P and the inferred regulation status in tumor.

| GENE | Log fold-change | | | | | Adj. $P$-val | Regulation Status |
| --- | --- | --- | --- | --- | --- | --- | --- |
| | Grade 1<br>$\beta_1$ | Grade 2<br>$\beta_2$ | Grade 3<br>$\beta_3$ | Grade 4<br>$\beta_4$ | Grade 5<br>$\beta_5$ | | |
| HOXC6 | 3.300889 | 3.599581 | 4.321112 | 4.13026 | 4.325551 | 1.25E-52 | UP |
| SIM2 | 3.063839 | 3.055925 | 2.992888 | 3.223932 | 2.958961 | 4.16E-51 | UP |
| EPHA10 | 2.054480 | 2.256922 | 2.353943 | 2.590398 | 2.337854 | 1.62E-50 | UP |
| HPN | 2.782889 | 2.830453 | 3.143309 | 3.049693 | 2.577042 | 7.29E-49 | UP |
| PYCR1 | 1.476672 | 1.524151 | 1.676698 | 1.669362 | 1.590206 | 3.37E-47 | UP |
| SLC19A1 | 1.723341 | 1.636842 | 1.660923 | 1.617484 | 1.484677 | 2.47E-46 | UP |
| TROAP | 1.27276 | 1.487522 | 1.897887 | 2.767342 | 2.943353 | 2.98E-46 | UP |
| LRFN1 | 1.622767 | 1.564225 | 1.686172 | 1.795652 | 1.786367 | 2.98E-46 | UP |
| EZH2 | 1.390663 | 1.442683 | 1.596514 | 2.098142 | 2.108934 | 1.04E-45 | UP |
| CGREF1 | 2.354448 | 2.340171 | 2.591191 | 2.508152 | 2.340949 | 7.61E-44 | UP |

To identify grade-specific genes, we binned the significant DE genes into partitions representing different grades, represented in Figure 4 as an UpSet plot [42]. To establish their salience, we identified genes that passed all the filter criteria described in the Methods section, and obtained ten grade I-salient, two grade II-salient, one grade III-salient, 34 grade IV-salient and 30 grade V-salient genes. The top 10 grade-salient genes in each grade are shown in Table 5; information on all the grade-salient genes is documented in Supplementary File S4. A heatmap of the grade-salient genes, shown in Figure 5, revealed a mostly systematic trend in their expression relative to the controls, with a mixture of up- and down-regulation processes. The map was clustered based on differential expression across grades and reflected a separation between high-grade and low-grade cancers. A dendrogram of the grade-salient genes yielded further insights into clustering among the grades, with a co-clustering among genes specific to indolent cancers (grades 1, 2, and 3) as well as among genes specific to aggressive cancers (grades 4 and 5). A volcano plot of the differentially expressed genes in PRAD is shown in Figure 6.

**Figure 4.**
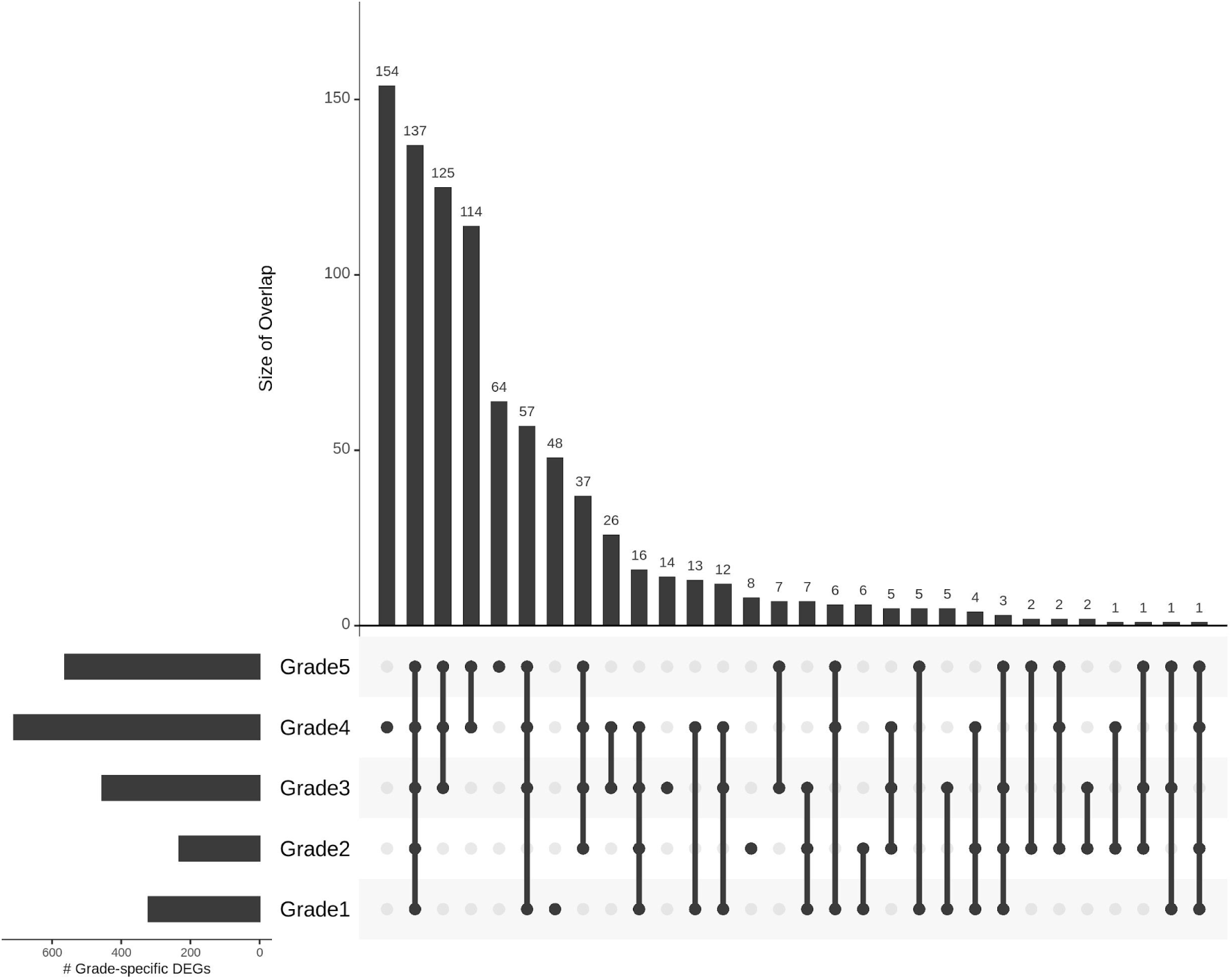
Gleason-grade distributions of the computationally identified grade-salient biomarkers, sorted by cardinality. An enrichment in the advanced grades is observable.

**Figure 5.**
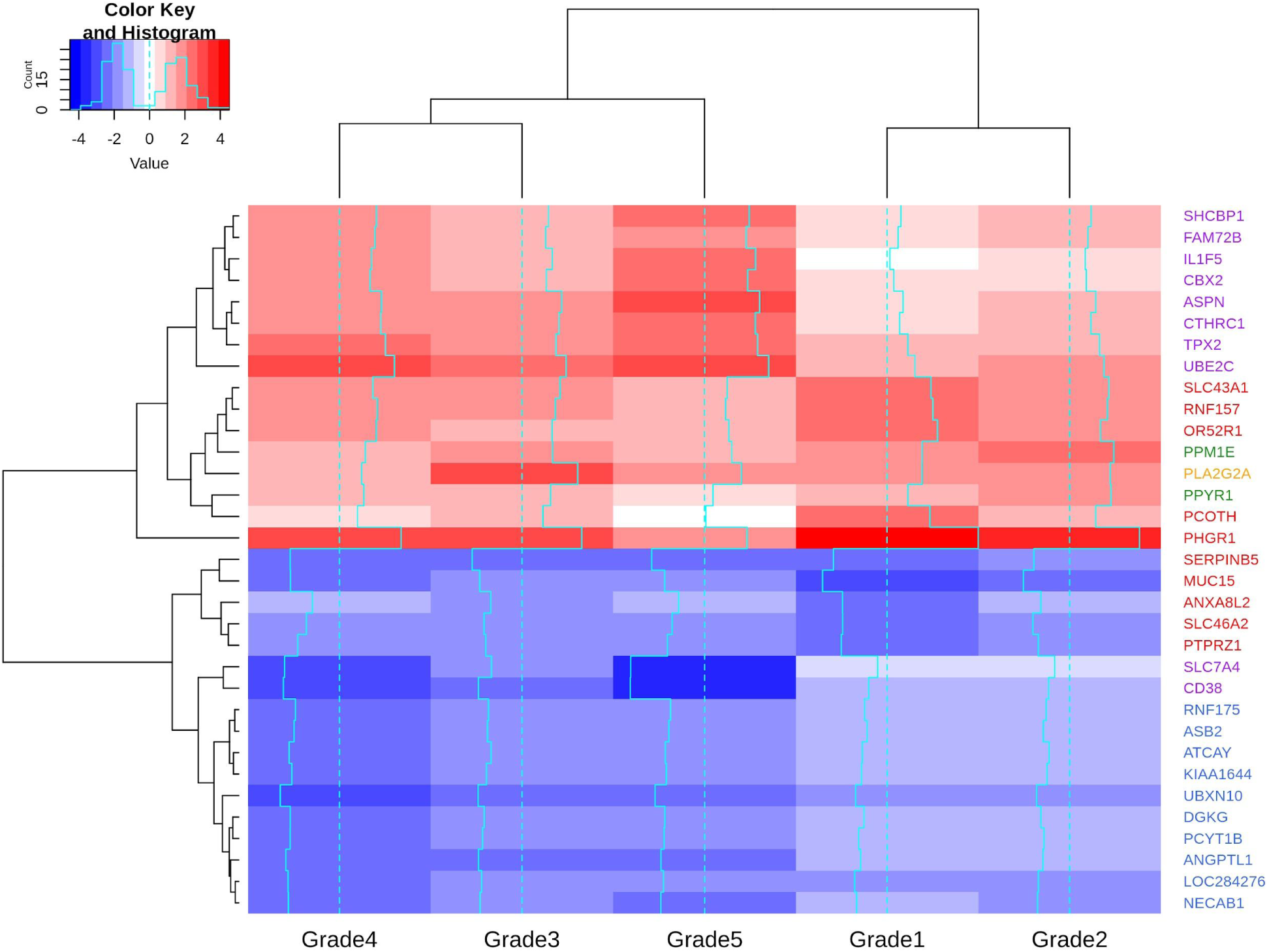
Heatmap of the expression of grade-salient genes, with clustering in both axes. Only the top ten grade-salient genes in each grade were used to construct the heatmap, for a total of 33 genes (as shown in Table 5). The downregulation of all the ten grade-4 salient genes is apparent.

**Figure 6.**
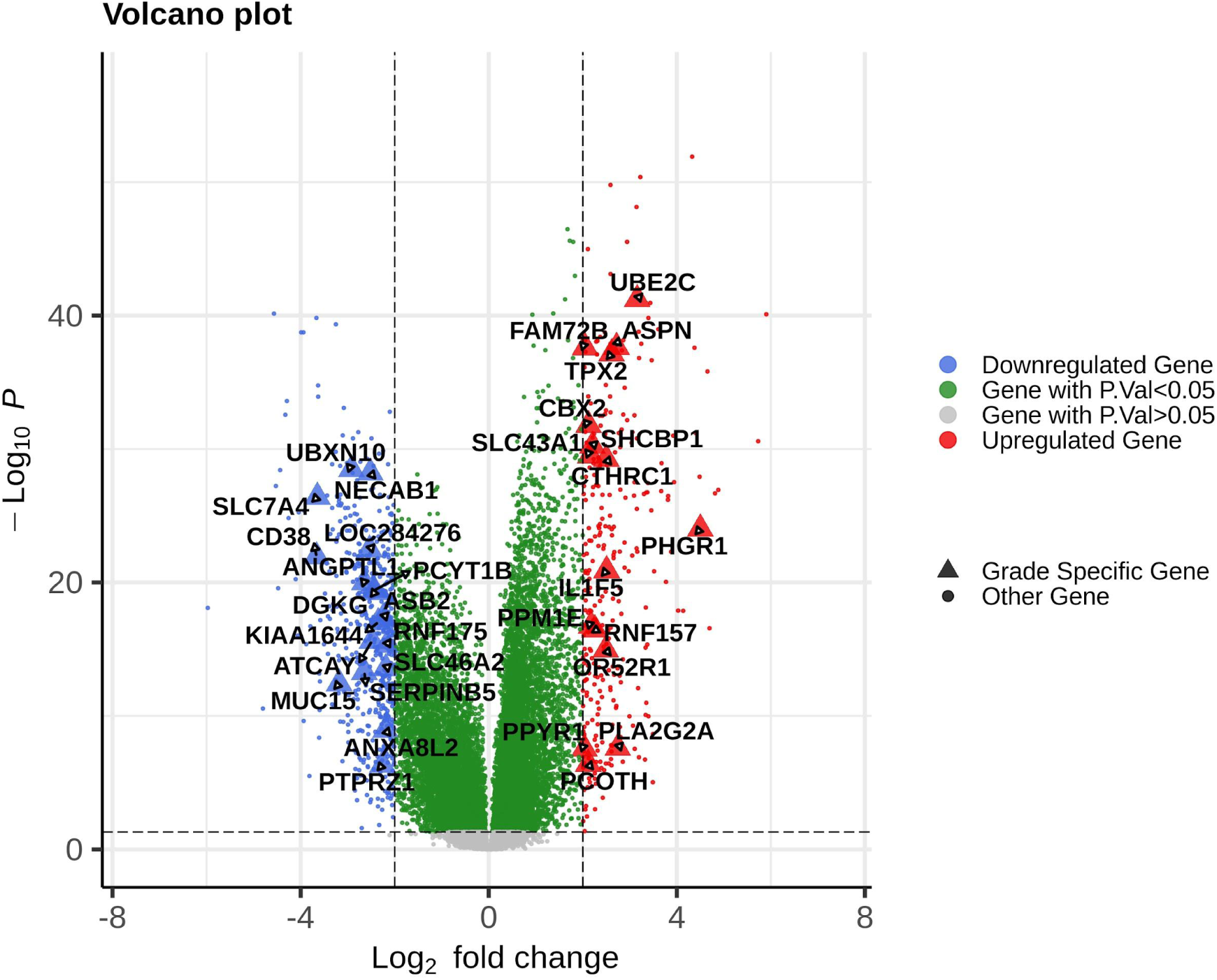
Volcano plot of the significant differentially expressed genes, with the grade-salient genes highlighted.

**Table 5:** The top grade-salient genes, by grade.

| Rank | Grade 1 | Grade 2 | Grade 3 | Grade 4 | Grade 5 |
| --- | --- | --- | --- | --- | --- |
| 1 | PHGR1 | PPM1E | PLA2G2A | UBXN10 | CBX2 |
| 2 | OR52R1 | PPYR1 |  | ATCAY | FAM72B |
| 3 | PCOTH |  |  | ANGPTL1 | SLC7A4 |
| 4 | SLC43A1 |  |  | DGKG | ASPN |
| 5 | ANXA8L2 |  |  | NECAB1 | UBE2C |
| 6 | RNF157 |  |  | ASB2 | SHCBP1 |
| 7 | MUC15 |  |  | KIAA1644 | TPX2 |
| 8 | SLC46A2 |  |  | LOC2824276 | IL1F5 |
| 9 | PTPRZ1 |  |  | PCYT1B | CTHRC1 |
| 10 | SERPINB5 |  |  | RNF175 | CD38 |

The genes in each grade are ranked by adj. p-values of the linear modeling analysis.

The top 200 genes from the numeric model (eqn. 3) are provided in Supplementary File S5. Applying the definition of monotonicity, we obtained 307 monotonically expressed genes, with 238 significantly monotonic (112 up and 126 down). The MEGs are also included in Supplementary File S5, ranked by significance. The intersection of significant ME genes with grade-specific genes yielded three genes: one grade-4 specific gene SERPINA5, and two grade-5 specific genes: DPYS and NKX6-1. Eleven genes were common to the top 200 genes from the numeric model and the significant MEGs, namely RCBTB2, VPS36, PLCD1, HRNBP3, NUPL2, SPAG5, SCRIB, AGL, CAMK2G, WHSC1, and ZNF706.

Gene set enrichment analysis using the grade-salient genes was performed on GO and KEGG to probe network effects. The grade-wise enrichment results are presented in Supplementary File S6. We screened the grade-salient genes against the CGC, but did not find any hits, suggesting that our findings could be novel to cancer biology, especially the Gleason-specified progression of prostate cancer. On screening the grade-salient genes against the NCG curated database of cancer drivers and healthy drivers, we found two hits, namely ANXA8L2 (grade-1 salient), and TPX2 (grade-5 salient). The annotation of TPX2 (‘putative oncogene’) agrees with the upregulation observed here. Searching the grade-salient genes in ClinicalTrials.gov, we found eight genes used as either targets or endpoints of clinical trials (Supplementary File S7). Three among these eight (namely MUC15, UBE2C, and CD38) were targeted in prostate cancer-related clinical trials, lending weight to our findings. The top 200 genes from linear model (eqn. 1) genes were screened against the Human Protein Atlas using the search: “cancer related genes” or “found as prognostic markers in other cancers” and the following hits were obtained: VAV2, BRAF, EZH2, MEN1, BUB1B, and MNX1. EZH2 and BRAF have been recently shown to promote castration-resistant prostate cancers [43,44], concordant with the upregulation in our analysis. VAV2 has been tied to poor prognosis in prostate cancer, corroborated by its oncogenic role observed here [45]. MEN1 has been noted as a tumor suppressor gene with protective effects in aggressive tumors, via increased gene expression in prostate cancer cells, thanks to inclusion in a high copy number gain region [46], which is corroborated by the upregulation in its expression here. Oncogenic BUB1B was necessary for fast proliferation of tumor cells [47], while oncogenic MNX1 has been associated with aggressive disease [48], both results supported by the upregulation found here. The monotonic grade-salient genes in the advanced aggressive stages of PRAD, namely DPYS, NKX6-1, and SERPINA5 have been documented in the prostate cancer literature. Aberrant hypermethylation of DPYS has been found to have prognostic value in stratifying early prostate cancers [49], discordant with the overexpression observed here. NKX6-1, observed here as an oncogene with respect to prostate adenocarcinoma, has been documented with opposing functional roles depending on cancer cell-type: as tumor suppressor in colorectal cancer, gastric cancer, acute lymphoblastic leukemia, B-cell lymphoma; and as oncogene in hepatocellular carcinoma and breast cancer [50]. Loss of expression of SERPINA5 is correlated with high-grade prostate tumors and adverse prognosis [51,52], concordant with the significant downregulation observed in the cancer samples here. The overlap of the top 200 monotonic genes with the top 200 genes from the linear model yielded three genes (SPAG5, ZNF706, SERPINA5), and with the top 200 of ordinal model (eqn. 3) yielded eleven genes: SPAG5, ZNF706, RCBTB2, VPS36, PLCD1, HRNBP3, NUPL2, SCRIB, AGL, CAMK2G, and WHSC1. A discussion of the grade-salient genes with corresponding violin plots is provided in Supplementary File S8 document.

### WGCNA Analysis

Grade-specific weighted co-expression networks were constructed using R WGCNA. The β parameter was identified as 14, 9, 7, 3, and 4 for Gleason grade-I, grade-II, grade-III, grade-IV, and grade-V networks, respectively. Identification of the optimal scale-free network is illustrated for grade-I in Figure 7. Modular decomposition was effected by the dynamic tree cut algorithm, and the outlier genes in each grade were binned into a gray module. This enabled the calculation of GS and MM of each gene (along with their *P*-values). This information is presented grade-wise in Supplementary File S9. Figure 8 shows the module-trait correlation for all the modules in each grade as a heatmap, with the source data given in Supplementary File S10.

**Figure 7.**
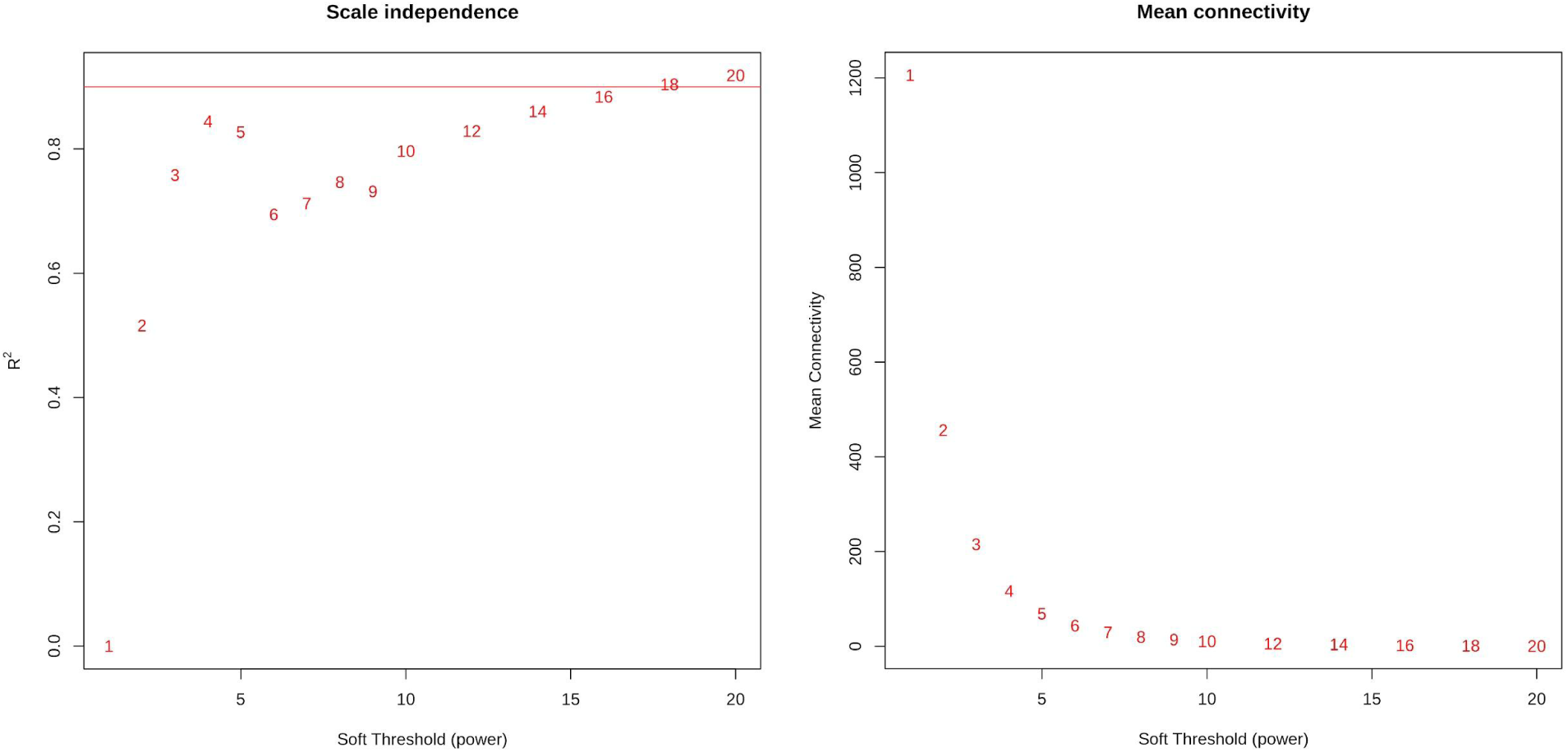
Determination of soft-thresholding power in WGCNA analysis, illustrated for Grade-I network. (A) Scale independence. Identifying the optimal soft-thresholding exponent β using the scale-free fit of degree distribution (i.e., *p(k)* ∼ *k*^−γ^) for various values of β. (B) Mean connectivity at different β.

**Figure 8.**
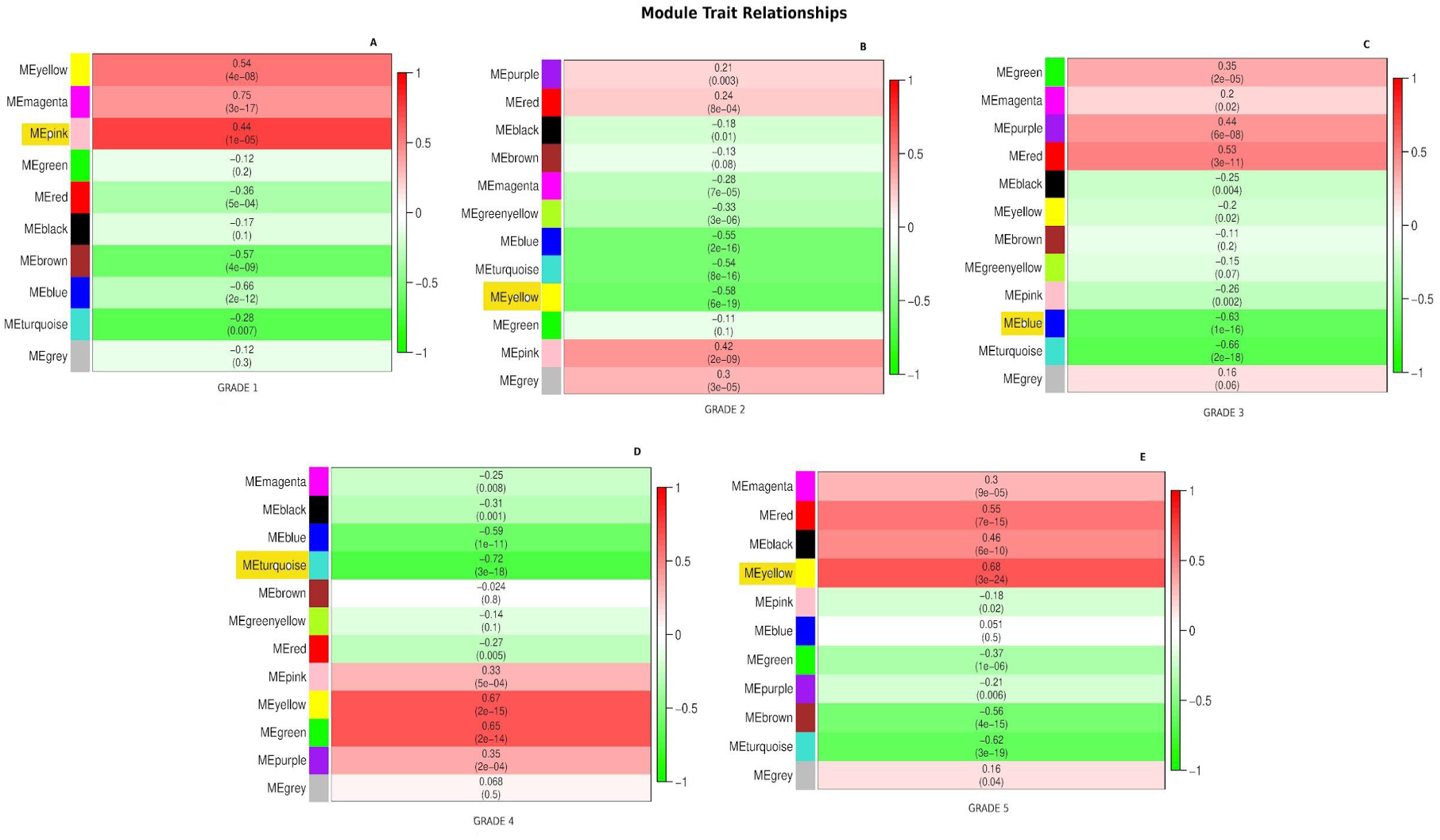
Heatmap of grade-wise module-trait relationships, depicting correlation (with *P*-value) between module eigengene (ME) and grade for each module in the given grade. (A) Grade-I, (B) Grade-II, (C) Grade-III, (D) Grade-IV, and (E) Grade-V. The Significant Module in each grade is highlighted. Strength of correlation is visualized using a color gradient.

Based on the per-module GS values, the Significant Module of each grade was identified, and a scatterplot of MM vs GS for all the Significant Modules was created (Figure 9). A strong Pearson’s correlation between MM and GS was observed for all Significant Modules, suggesting intrinsic importance for the respective grades. The trait-specific key genes in each Significant Module were identified, obtaining 67 genes in Grade-I, 46 genes in Grade-II, 204 genes in Grade-III, 603 genes in Grade-IV, and 83 genes in Grade-V represented in Table 6.

**Figure 9.**
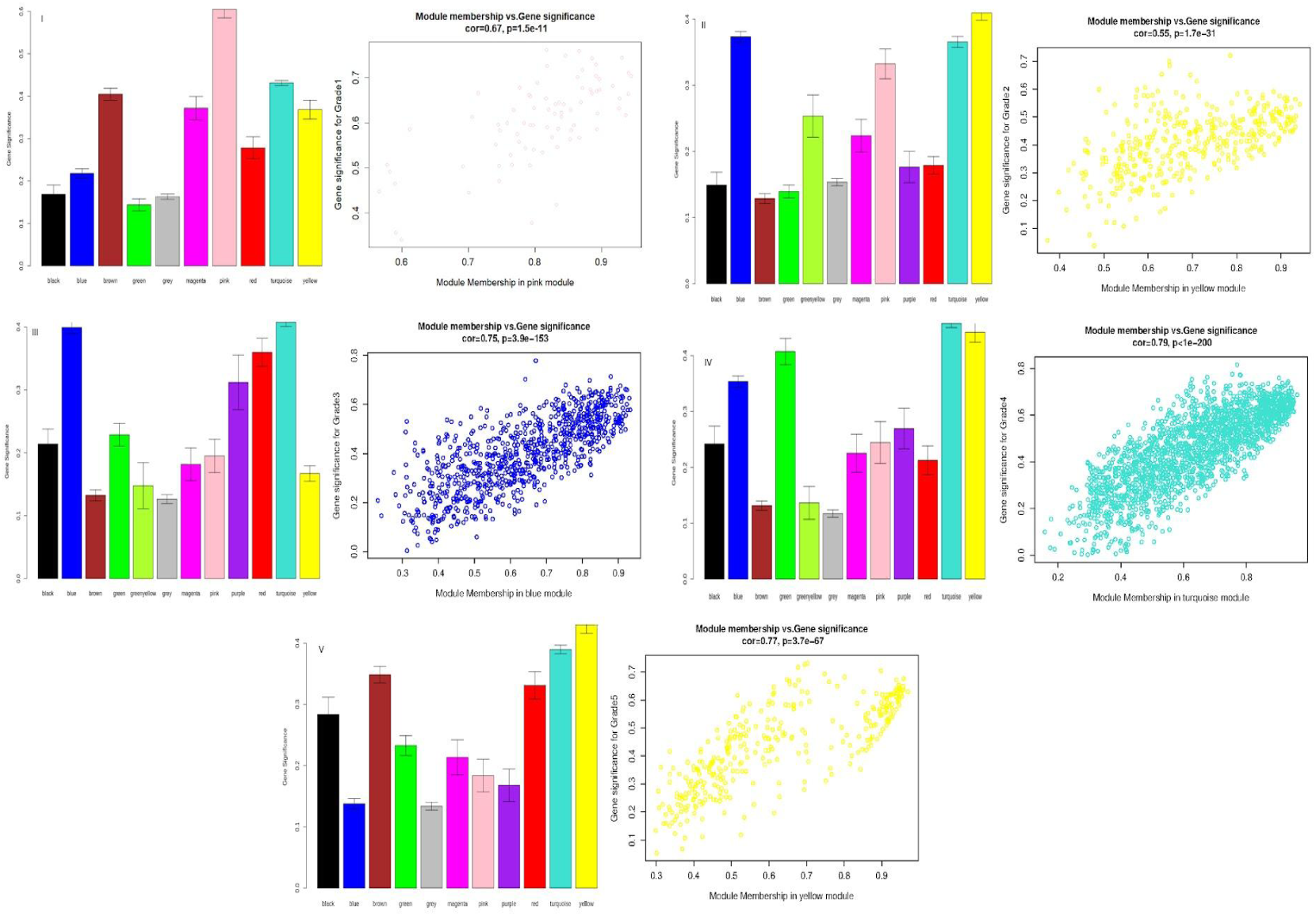
Grade-wise module significance, along with the scatter of genewise correlation between MM and GS in the grade’s Significant Module. (I) Grade-I; (II) Grade-II; (III) Grade-III; (IV) Grade-IV; and (V) Grade-V.

**Table 6:** Summary of Gleason grade-wise WGCNA analysis of PRAD transcriptome.

| | $\beta$ | No. of Modules | Significant Module | # Genes in Sig. Module | Key Genes in Sig. Module | ME-Trait correlation | Significance of trait corr. ($P$) |
| --- | --- | --- | --- | --- | --- | --- | --- |
| Grade 1 | 14 | 10 | Pink | 79 | 67 | 0.44 | 1e-05 |
| Grade 2 | 9 | 12 | Yellow | 381 | 46 | -0.58 | 6e-19 |
| Grade 3 | 7 | 12 | Blue | 843 | 204 | -0.63 | 1e-16 |
| Grade 4 | 3 | 12 | Turquoise | 1879 | 603 | -0.72 | 3e-18 |
| Grade 5 | 4 | 11 | Yellow | 336 | 83 | 0.68 | 3e-24 |

The top ten trait-specific key genes in the significant module in each grade were used to reconstruct driver networks using STRINGdb. The results are detailed in Supplementary File S11. The trait-specific key genes of Grade-1 WGCNA network were enriched in the catabolic processes of fatty acids and lipids (*P* < 1E-04). The trait-specific key genes of Grade-2 WGCNA network showed a KEGG enrichment of PPAR signaling pathway (*P* < 1E-2), altering androgen activity thereby driving PRAD progression [53]. The trait-specific key genes of Grade-3 WGCNA network revealed a Reactome enrichment [54] for neddylation pathway (*P* < 1E-04), which increases androgen receptor transcription and promotes the growth and invasion of the prostate cancer cells [55]. The trait-specific key genes of Grade-4 WGCNA network showed an enrichment in post-translational SUMOylation modifications in the pTEN/AKT and androgen-receptor signaling pathways (*P* < 2E-3) [56]. An analysis with Reactome revealed enrichment in PPARA gene expression (*P* < 0.05), known to drive advanced prostate cancer [57]. The top ten trait-specific key genes of Grade-5 WGCNA network were enriched in pTEN transcription regulation (*P* < 1E-04), whose loss of function is well-known to be associated with aggressive and metastatic prostate cancer [58]. An analysis with Reactome showed significance for HCMV early events (*P* < 1E-3), known to be involved in the etiology of metastatic prostate carcinoma [59].

## DISCUSSION

We have executed independent workflows for the statistical and network-based modeling of the TCGA PRAD transcriptome, stratified by Gleason grade. Based on the outcomes of these workflows, we investigated the areas of concurrence between the results that would represent consensus and robust findings. The following observations were notable:

i. All the grade-salient genes were located either in the significant modules or with a module highly correlated with the grade;
ii. Grade-salient genes displaying significant upregulation with Gleason progression almost always yielded a significant positive GS in the WGCNA analysis, except one: MAL;
iii. Grade-salient genes displaying significant downregulation with Gleason progression almost always yielded a significant negative GS in the WGCNA analysis, except four: COL10A1, NOX4, FAP, and SFRP4.
iv. Regulation status of trait-specific key genes suggested by the sign of the GS was concordant with the inference from the statistical expression patterns with respect to controls, with one exception: LOC100128675.
v. All the grade-salient genes invariably reflected a strong and significant association with their WGCNA modules in the respective grades (MM > 0.4, *P* < 0.05).

The results of the consensus analysis are detailed in Supplementary File S12, and encouraged the investigation of deeper concordance between the two orthogonal computational approaches. The grade-wise overlap between grade-salient genes from the statistical modeling and the trait-specific key genes from network modeling included two genes from Grade-1 (SLC43A1, PHGR1), 26 genes from Grade-4 (including C2orf88, ANGPT1, CAV2, TMLHE, IGSF1, PPARGC1A, LGR6, PPP1R3C, FRMD6, NECAB1; please see Supplementary File S12) and seven genes from Grade-5 (CBX2, FAM72B, SHCBP1, TMEM132A, TPX2, DPYS and UBE2C). The regulation status for these 35 consensus genes was concordant, underscoring their potential utility. A summary of the consensus genes is presented in Table 7. A gene-by-gene analysis of the trait-specific top ten key genes in each module is presented in Supplementary File S13. The main inference remained unaltered: the bulk of the trait-specific key genes were also grade-salient, with sync in inferred regulation. It is striking that most of the consensus emerged from the aggressive forms of prostate cancer (i.e. Gleason grades 4 and 5). We used the 33 consensus genes from the aggressive grades to reconstruct a STRINGdb network, with 50 interactors in the first shell and 10 interactors in the second shell. This yielded a significantly enriched PPI with 542 edges (P < 1.0E-16; Figure 10). An analysis with KEGG revealed significant enrichment in oncogenic pathways like NF-kappa B signaling pathway (P < 1E-3), and p53 signaling pathway (P ∼ 0.02). An analysis with Reactome showed significant enrichment in cell cycle processes. The detailed results of these analyses are presented in Supplementary File S14 table. The consistency in the results between the –omics analysis and WGCNA is complete, which substantively amplifies the significance of the findings and sets the stage for modeling the character (aggressive or not) of PRAD samples based on these results.

**Figure 10.**
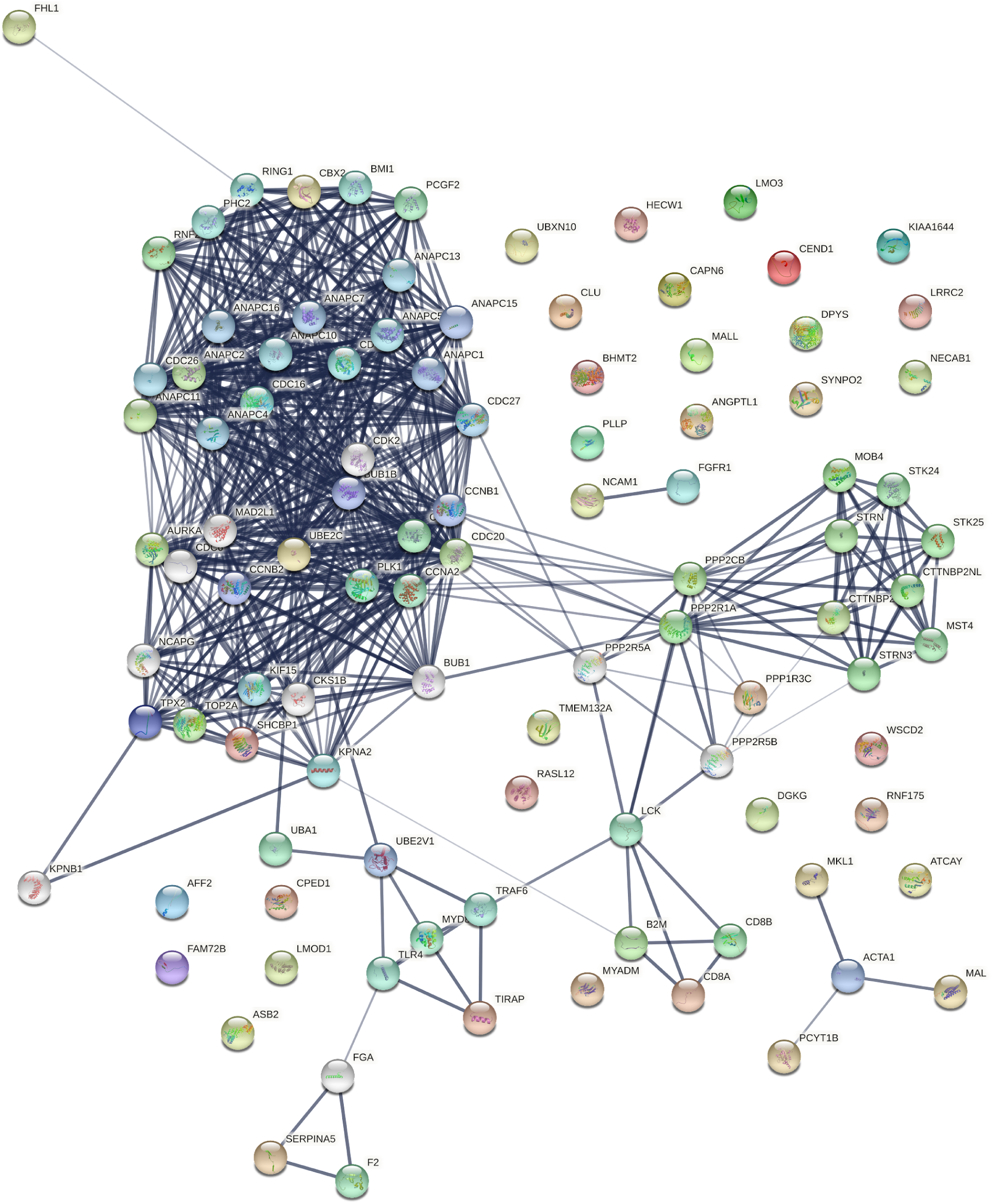
Network reconstructed using the 33 consensus root biomarkers of the aggressive grades of prostate cancer. A giant component with a clique-like core could be seen. It is remarkable that 25 of the consensus root biomarkers were isolated outliers, signifying their roles underpinning varied biological processes not immediately related to each other.

**Table 7:**
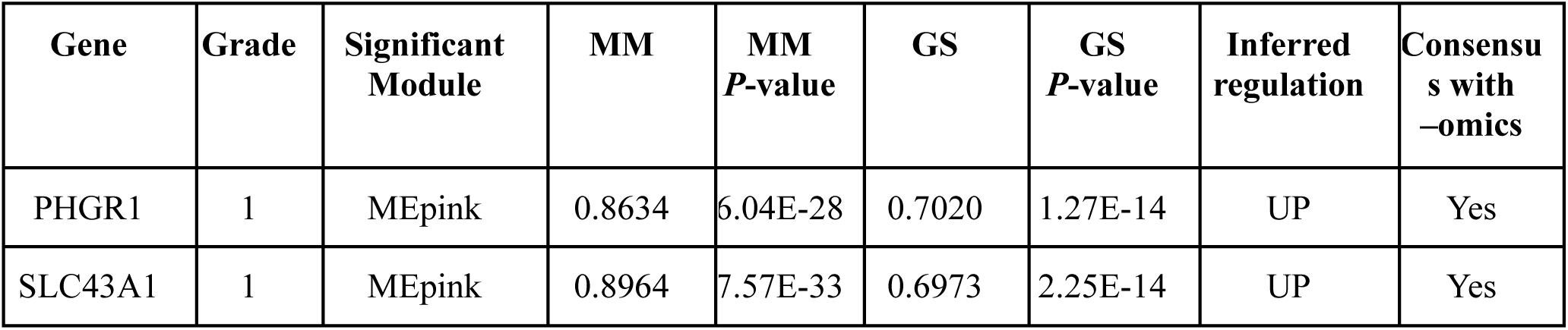
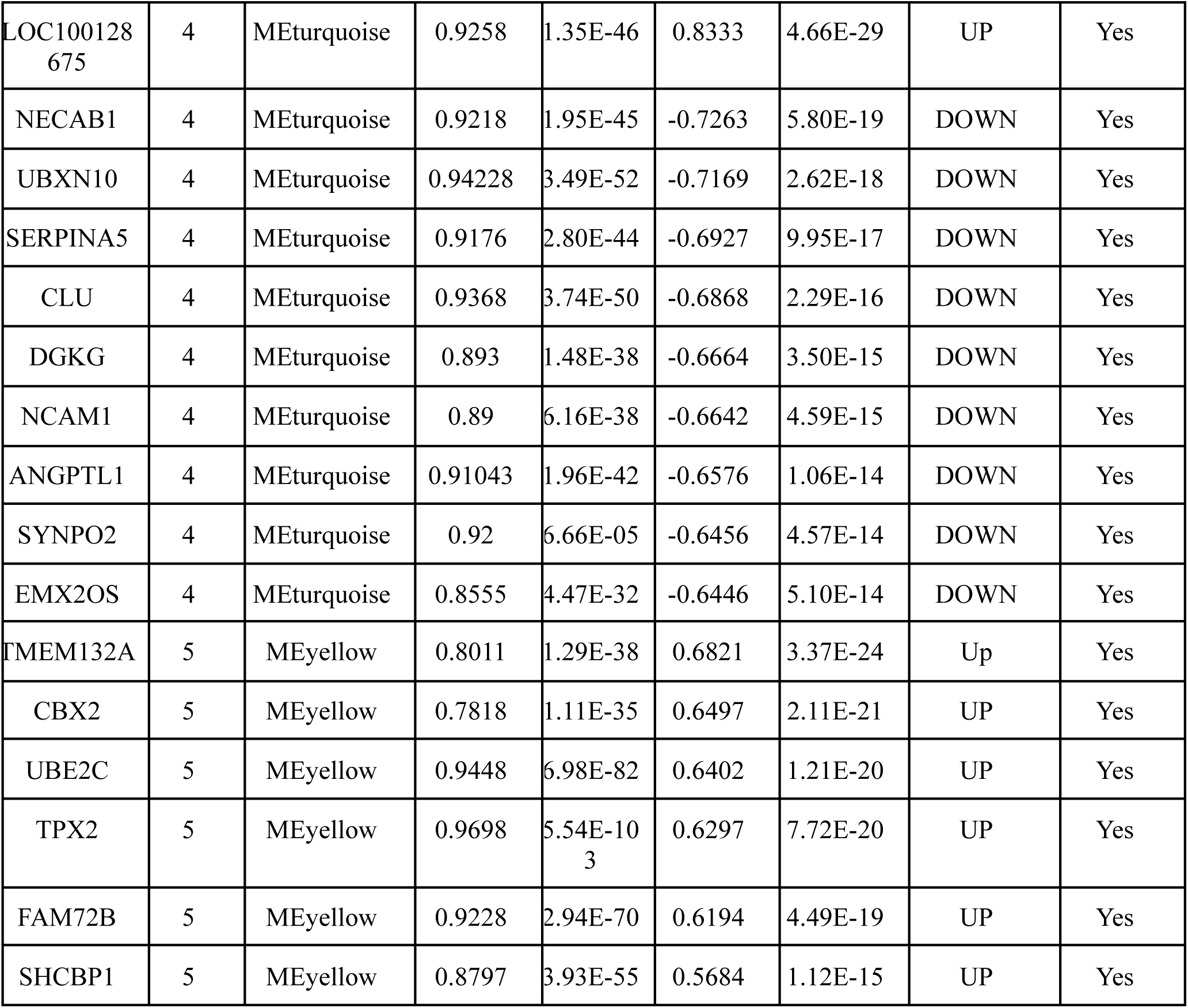
Grade-wise root biomarkers from consensus analysis of grade-salient genes from statistical analysis and trait-specific key genes from network analysis. In all cases, the Significant Module for the grade matches the module with which the gene has the largest MM. The inferred regulation is based on the sign of the GS, which denotes the correlation between gene expression and trait class (Gleason grade) of interest. Only the top ten genes from Grade-4 (ranked by GS) are shown. Consensus in regulation inferred from statistical analysis is also reported, and seen to be concordant throughout.

The expression subset of the foregoing 35 consensus genes were the features, and the 538 samples were the instances for developing ML models for pre-screening PRAD and typing PRAD aggressiveness. As noted in Methods, the samples were annotated as benign, indolent or aggressive, and the mapping between this outcome and the feature space was modeled using various classifiers (Table 8). Hyperparameters were optimized using 10-fold cross-validation. Different kernels were tried for the SVM classifier (viz. linear, radial and polynomial), and results for only the kernel with the best performance are shown. The RandomForest model with optimal hyperparameters showed a balanced accuracy ∼ 100.00% among the three classes of interest during cross-validation. SVM with radial kernel and XGBoost were also effective models, with >90% cross-validated balanced accuracy. We investigated the RandomForest model with perfect discrimination for feature importance using R caret [60] (Figure 11). The top ten features based on mean decrease in Gini score [61] were SLC43A1, FAM72B, CBX2, UBE2C, SHCBP1, TMEM132A, LOC100128675, TPX2, PHGR1, DPYS. Both the consensus grade-I salient genes (SLC43A1 and PHGR1) were key features in the RandomForest solution, as were all the seven consensus grade-V salient genes (FAM72B, CBX2, UBE2C, SHCBP1, TMEM132A, TPX2, and DPYS), validating the significant contribution of differential Gleason grade-specific biomarkers to the aggressive character of PRAD cancers.

**Figure 11.**
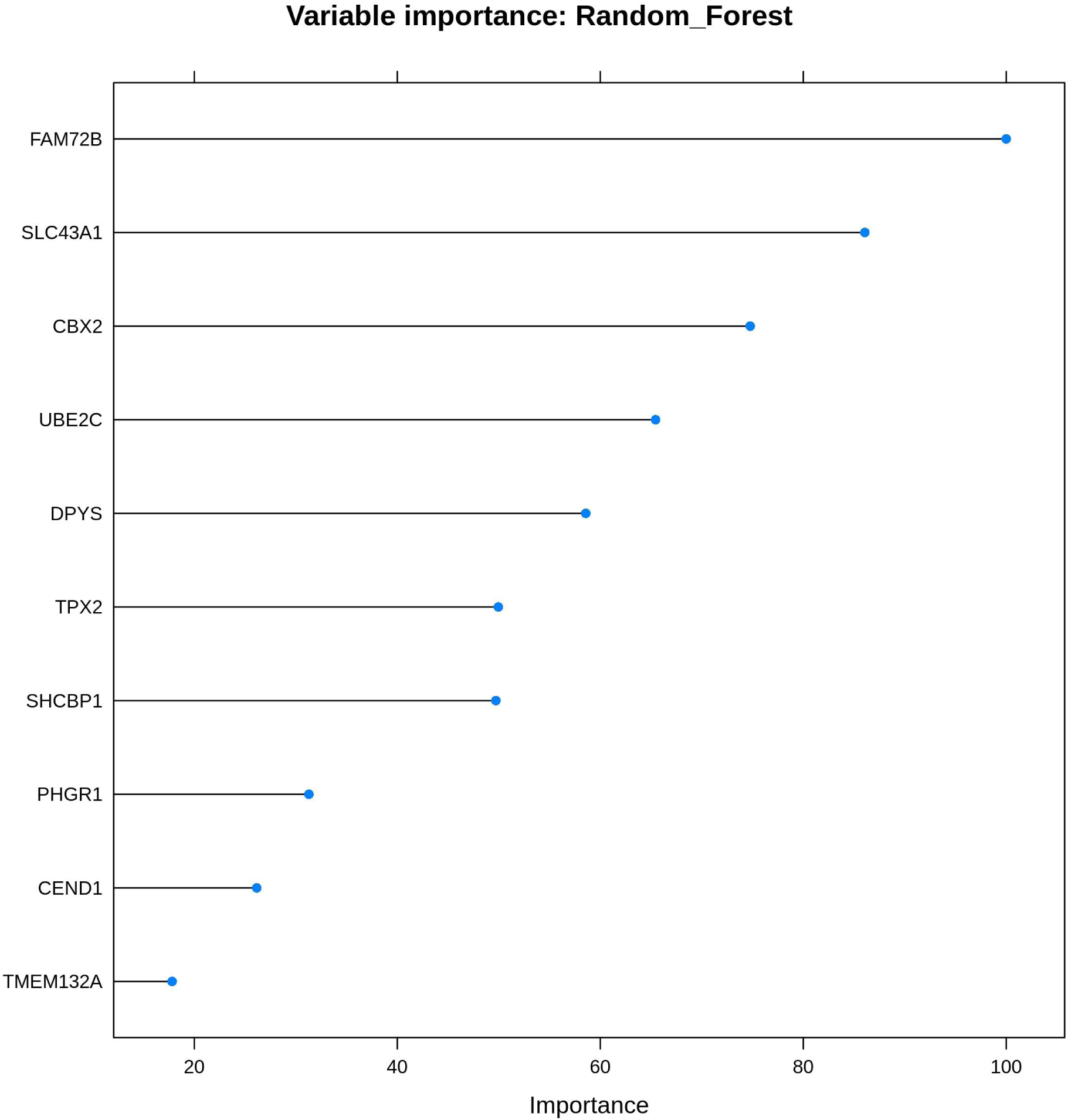
Elucidation of feature importance for the best-performing RandomForest model. Shown are the top ten features enabling the detection of prostate cancers with aggressive character. SLC43A1, a Gleason grade-I root biomarker, has the greatest contribution to the classification problem, followed by many Gleason grade-V root biomarkers (FAM72B, CBX2, UBE2C, SHCBP1, TMEM132A). Also seen is LOC100128675, the lone Gleason grade-IV root biomarker among 26 such features.

**Table 8:** Hyperparameters and performance measures of different models investigated for the ternary classification problem of sample as normal (possibly benign hyperplasia), indolent prostate cancer or aggressive prostate cancer, based on expression levels of root consensus genes. Optimal values of hyperparameters are provided in the same order as the hyperparameters considered. Performance was measured using 10-fold cross-validation balanced accuracy for the multi-class problem.

| S.No | Classifier | Hyperparameters of interest | Optimal hyperparameters | CV balanced accuracy (%) |
| --- | --- | --- | --- | --- |
| 1 | SVM (radial kernel) | cost, gamma | 1, 0.1 | 93.87 |
| 2 | RandomForest | ntree (# trees in the forest), mtry (# candidate variables randomly sampled for splitting) | 500, 6 | 100.00 |
| 3 | Neural Networks (1-layer) | size, decay | 2, 1 | 77.94 |
| 4 | Neural Networks (2-layer) | #units in hidden layer 1, #units in hidden layer 2 | 2, 26 | 89.18 |
| 5 | XGBoost | eta, max depth, colsample_bytree | 0.01,5,1 | 92.37 |

The better-performing models, viz. RandomForest, SVM (radial kernel) and XGBoost, were re-built using the full dataset, and these are provided in Supplementary File S15 as RDS binaries for academic and not-for-profit use. The hyperparameter-optimized best-performing RandomForest model, PRADclass, has been deployed as a web-server (https://apalania.shinyapps.io/pradclass/) to facilitate the pre-screening of PRAD cancers and typing their aggressive character. Features from expression profiling have been earlier used to build predictive models of cancers, for e.g. breast cancer [62]. PRADclass may assist in risk stratification independent of deep learning systems, especially in settings where availability of Gleason grading expertise is constrained, and necessitates prospective clinical evaluation. It is a handy, documented, and readily available alternative decision support tool. The grade-wise consensus root biomarkers could indicate potential targets for therapy, suggesting experimental investigations. Future research may address the fine discrimination among Gleason grades.

## CONCLUSION

Gleason grading is the gold standard for risk stratification of prostate cancer patients, and molecular biomarkers specific to different Gleason grades could enable the diagnosis of high-risk prostate cancers requiring active management. In this study, we have applied a couple of orthogonal computational pipelines, based on statistical modeling and network analysis of expression data, to identify grade-wise consensus root biomarkers. Statistical modeling yielded ten grade-I salient genes (PHGR1, OR52R1, PCOTH, SLC43A1, ANXA8L2, RNF157, MUC15, SLC46A2, PTPRZ1, and SERPINB5), two grade-II salient genes (PPM1E and PPYR1), one grade-III salient gene including (PLA2G2A), 34 grade-IV salient genes (including UBXN10, ATCAY, ANGPTL1, DGKG, NECAB1, ASB2, KIAA1644, LOC2824276, PCYT1B, RNF175), and 30 grade-V salient genes (including CBX2, FAM72B, SLC7A4, ASPN, UBE2C, SHCBP1, TPX2, IL1F5, CTHRC1, CD38). Post WGCNA modeling, the consensus analysis yielded 35 root biomarkers, viz. two genes in Grade 1 (SLC43A1, PHGR1), 26 genes in Grade 4 (LOC100128675, PPP1R3C, NECAB1, UBXN10, SERPINA5, CLU, RASL12, DGKG, FHL1, NCAM1), and seven genes in Grade 5 (CBX2, DPYS, FAM72B, SHCBP1, TMEM132A, TPX2, UBE2C). These biomarkers might double as novel chemotherapy target hypotheses, necessitating experimental corroboration. To explore the histoprognostic value of these molecular markers, we constructed various machine learning models for the ternary problem of classifying a sample as non-cancerous, indolent prostate cancer or aggressive prostate cancer. A RandomForest model, PRADclass, secured a cross-validated balanced accuracy ∼ 100.00%, indicating potential uses for cancer pre-screening and character typing. PRADclass is available on the web for academic and non-commercial use at https://apalania.shinyapps.io/pradclass. Further work is necessary to assess the clinical utility of PRADclass as a readily available decision support aid in the treatment-management of malignant prostate gland neoplasia.

## Data Availability

All data produced are available online at:
https://doi.org/10.6084/m9.figshare.22549621.v1

## Abbreviations

CGC: Cancer Gene Census
CV: cross-validation
DEG: Differentially expressed genes
DRE: Digital rectal examination
GO: Gene Ontology
GS: Gene significance
KEGG: Kyoto Encyclopaedia of Genes and Genomes
ISUP: International Society of Urological Pathology
lfc: log fold-change
ME: Module eigengene
MEG: Monotonically expressed genes
MM: Module membership
MS: Module significance
NCG: Network of Cancer Genes
PCA: Principal Components Analysis
PRAD: Prostate adenocarcinoma
PSA: prostate-specific antigen
RSEM: RNA-Seq by Expectation Maximization
TCGA: The Cancer Genome Atlas
TNM: Tumor, Node, Metastasis
TOM: Topological overlap matrix
SVM: Support Vector Machine
WGCNA: Weighted gene co-expression network analysis

## Acknowledgements

We would like to thank Amrutha Karthikeyan and R. Shivathmika for assistance with the WGCNA analysis. We are grateful to the School of Chemical and Biotechnology, SASTRA Deemed University, for infrastructure and computing support.

## Data availability

All supplementary materials are available at: https://doi.org/10.6084/m9.figshare.22549621.

